# Biological mechanisms underlying the link between social adversity and cognition: A scoping review

**DOI:** 10.1101/2025.05.14.25327640

**Authors:** A Liang, E Watt, N Gomaa

## Abstract

**Background:** Various studies have shown that social adversity, such as loneliness or low SES, are linked with worse cognitive outcomes, though underlying biological mechanisms remain unclear. This scoping review aims to summarize existing evidence on biological processes that may underline this association.

**Methods:** Following PRISMA-ScR guidelines, studies measuring social adversity, cognition, and at least one biological mechanism were included. Results were summarized narratively and in tabular formats.

**Results:** Thirteen studies examined links between social adversity, cognition, and biological mechanisms. Inflammation, allostatic load, genetics and genetic aging markers were identified as potential mediators.

**Conclusion:** Several studies suggest that these biological mechanisms may mediate the link between social adversity and cognitive decline. However, further research is needed to clarify these complex relationships, which are crucial for developing targeted interventions, especially for socially disadvantaged populations.

## 1. Introduction

Aging is a multifaceted process that encompasses physical, cognitive, and emotional changes. As the global population continues to age, understanding the factors that influence cognitive health in later life has become a critical area of research. Increasing evidence highlights the significant role of social exposures, such as social engagement, socioeconomic status (SES), social support, and social networks, in shaping cognitive trajectories during aging(1,2). For an example, studies have consistently shown that social engagement is protective against cognitive decline, with older adults who maintain active social lives exhibiting better cognitive performance and a reduced risk of dementia(1,3). Conversely, social isolation and loneliness have been linked to an increased risk of cognitive impairment and dementia(3,4). In a longitudinal study, Wilson and colleagues found that individuals with higher levels of social activity were less likely to develop Alzheimer’s disease, suggesting that cognitive stimulation from social interactions may help preserve brain function in aging(5). The association between social networks and cognitive health is further supported by studies showing individuals with broader and more supportive social ties to experience slower rates of cognitive decline(2,6). In the coming decades, older adults are expected to make up a third of Canada’s population, and cognitive impairments can have profound negative impacts on an older adult’s life(7). One cohort study showed associations between mid-life marital status and later cognitive functions, where being divorced or widowed were both associated with greater odds of later cognitive impairments(8). Another cohort study showed social isolation and loneliness in older adults are both associated with lower cognitive scores(9). SES also plays a critical role in cognitive health during aging. Low SES, characterized by limited access to resources, education, and healthcare, has been consistently associated with worse cognitive outcomes and a higher risk of dementia(3). The impact of SES on cognitive aging is postulated to be mediated through various pathways, including chronic stress, limited access to healthcare, and less cognitive stimulation throughout life(10). These findings underscore the importance of social environments and experiences in shaping cognitive trajectories during aging, pointing to the need for integrated approaches that consider both individual and social factors. However, despite the wealth of evidence supporting the link between social exposures and cognitive outcomes in older adults, the mechanisms that underlie these associations remain unclear.

The biopsychosocial model of health, which was first proposed by Engel in 1977, emphasizes the interaction between biological, psychological, and social factors in influencing health outcomes(11,12). In the context of cognitive aging, this model provides a comprehensive framework for understanding how social exposures contribute to cognitive function. Social factors, such as social support and engagement, interact with psychological and biological processes to influence cognitive outcomes. For instance, psychological factors like stress, coping strategies, and mental health are known to interact with social support to affect cognitive health(13). Chronic stress, often exacerbated by adverse social environments, has been implicated in cognitive decline through its effects on the hypothalamic-pituitary-adrenal (HPA) axis, leading to elevated levels of cortisol that can negatively impact brain regions such as the hippocampus, which is crucial for memory and learning(14). Moreover, social engagement may buffer against the negative effects of stress by promoting positive psychological states, such as a sense of belonging and self-worth, which, in turn, may protect against cognitive decline(13,15). At the biological level, several mechanisms have been proposed to explain how social exposures may influence cognitive aging. One of the most studied mechanisms is the role of inflammation. Chronic stress and social adversity have been shown to increase inflammatory markers, which in turn may contribute to neurodegenerative processes and cognitive decline(16,17). Inflammatory responses, particularly the activation of microglia and the release of proinflammatory cytokines, have been linked to brain aging and Alzheimer’s disease pathology(18). Another potential biological pathway is neurotrophic support. Social engagement has been shown to enhance the release of brain-derived neurotrophic factor (BDNF), a protein crucial for neuronal survival and plasticity, thereby promoting cognitive resilience in the face of aging(19). Another potential mechanism linking social exposures to cognitive health in older age is through epigenetics(20,21). Studies over the last decade have shown that age-related epigenetic changes quantified in epigenome-wide association studies or those characterizing the epigenetic clock are linked to cognitive health in older age and may be modified by social exposures(13). Other mechanisms that have been explored include increased allostatic load, which is defined as the "wear and tear" the body experiences when repeated neural or neuroendocrine responses are activated during stressful situations, as well as telomere shortening(14,22–24).

This scoping review aims to summarize the existing evidence on the association between measures of social adversity and neurocognitive outcomes in adults and describe the underlying biological processes and pathways. By mapping the existing evidence and identifying key research areas, we hope to contribute to a deeper understanding of how social, psychological, and biological factors converge to influence cognitive outcomes in later life. This knowledge is crucial not only for advancing the science of cognitive aging but also for developing targeted interventions that address the social determinants of cognitive health and that promote healthy aging.

## 2. Methods

This manuscript was developed in accordance with the Preferred Reporting Items for Systematic Reviews and Meta-Analyses extension for Scoping Reviews (PRISMA-ScR) Checklist(25). A review protocol can be accessed on OSF registries (DOI 10.17605/OSF.IO/2DFWM).

### 2.1 Eligibility criteria

Eligible studies examined human adult populations and included measures of social adversity as exposures, measures of cognitive outcomes, and measures of biological mechanisms that link exposure and outcome. Social adversity is defined according to the WHO Commission on the Social Determinants of Health Framework which includes income and social protection, education, unemployment and job insecurity, working life conditions, food insecurity, housing, basic amenities, and environment, social inclusions and non-discrimination(26). We did not include early childhood development. Cognitive measures included any studies who assessed either general cognition or a specific cognitive domain(3,27). All study designs were included. Animal studies, and those that focused on pediatric populations, and studies not in the English language were excluded. We also excluded the grey literature, editorials, and other reviews or narrative articles.

### 2.2 Data sources and search strategy

A research librarian at Western University libraries was consulted in the development of the search strategy. Relevant documents were identified through literature searches conducted in databases MEDLINE, Embase, PsychINFO (all via Ovid interface), and Scopus. Keywords and medical subject headings (MeSH) were used to identify relevant studies, and no date restrictions were placed on the searches. Manual citation screening of included trials was also conducted to ensure a comprehensive search. The complete search strategy is available in the Supplementary Appendix (S1 Appendix).

### 2.3 Study selection and screening

All literature search results were uploaded to Covidence software and duplicates were removed through an automated duplication check conducted by Covidence and a manual duplicate check conducted by two reviewers (AL and EW). The two reviewers (AL and EW) then independently conducted a title abstract screening of the eligible retrieved articles using Covidence. Potentially relevant studies then underwent full text screening by both reviewers independently. After each level of screening, conflicts were resolved during a consensus meeting between the two reviewers. Cohen’s kappa (K) coefficient was calculated for each level of screening to assess for inter-rater reliability in screening obtaining a coefficient range of 0.68-0.81 indicating substantial agreement between the reviewers.

### 2.4 Data charting and synthesis

Data was extracted from the included studies independently by the two reviewers. Extracted data pertained to study characteristics (authors, publication date, study design, location, number of participants), study variables (social adversity measure(s), cognition measure(s), biological mechanism measure(s)), and study outcomes (associations between studied variables).

### 2.5 Evidence appraisal and data synthesis

The Newcastle-Ottawa Scale was used to assess the risk of bias and quality (12). Completed risk-of-bias table is in the Supplementary Appendix (S2 Appendix). To summarize results of our data extraction, data from the included studies were summarized in a tabular format.

A narrative summary of the results was also provided in line with the Synthesis Without Meta-analysis (SWiM) guidelines and following the PRISMA-ScR checklist (13, 14).

## 3. Results

The overall results of the study are summarized in the visual abstract (Figure 1).

**Figure 1.**
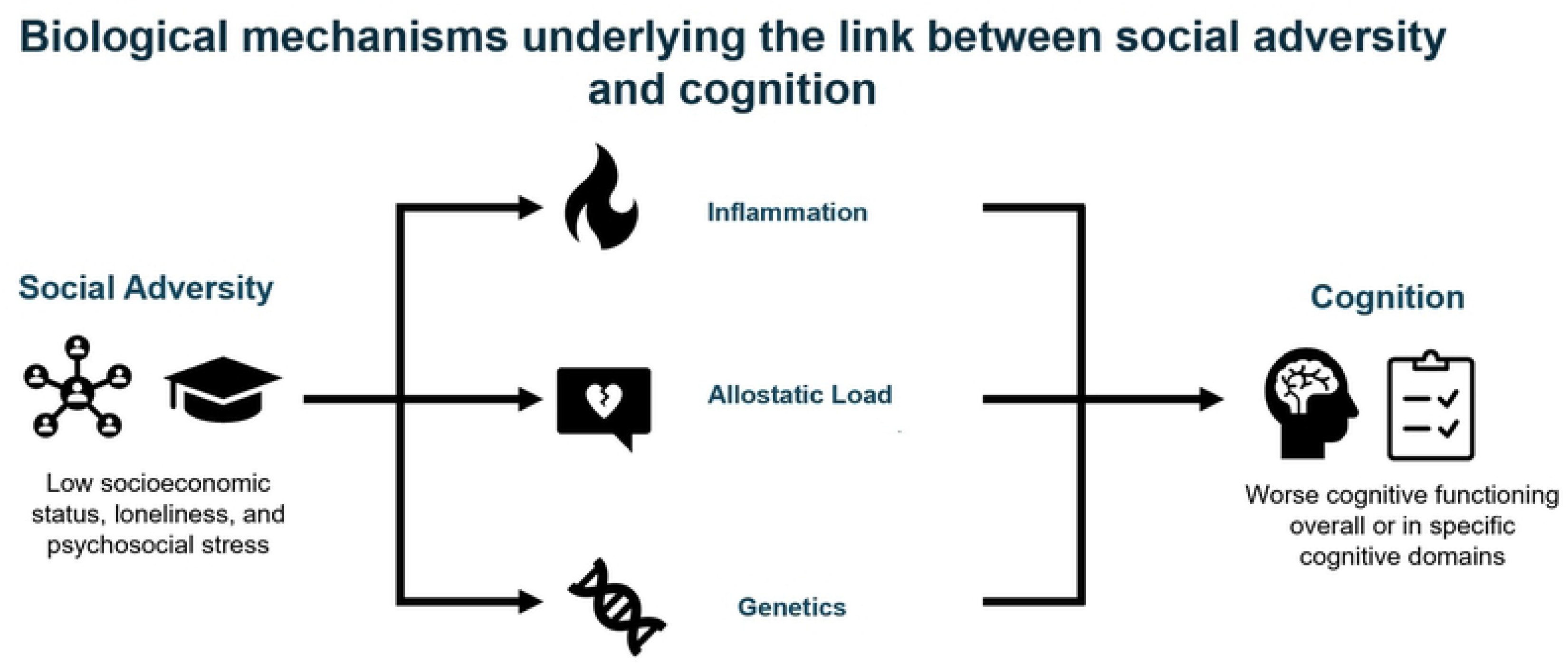
Visual abstract

### 3.1 Study selection and characteristics

The search strategy yielded 1872 results. After title and abstract screening, 207 studies remained and underwent full-text screening, out of which 13 studies met the inclusion(13,28–39). Figure 2 summarizes the evidence of the studies screened and exclusions at each stage.

**Figure 2.**
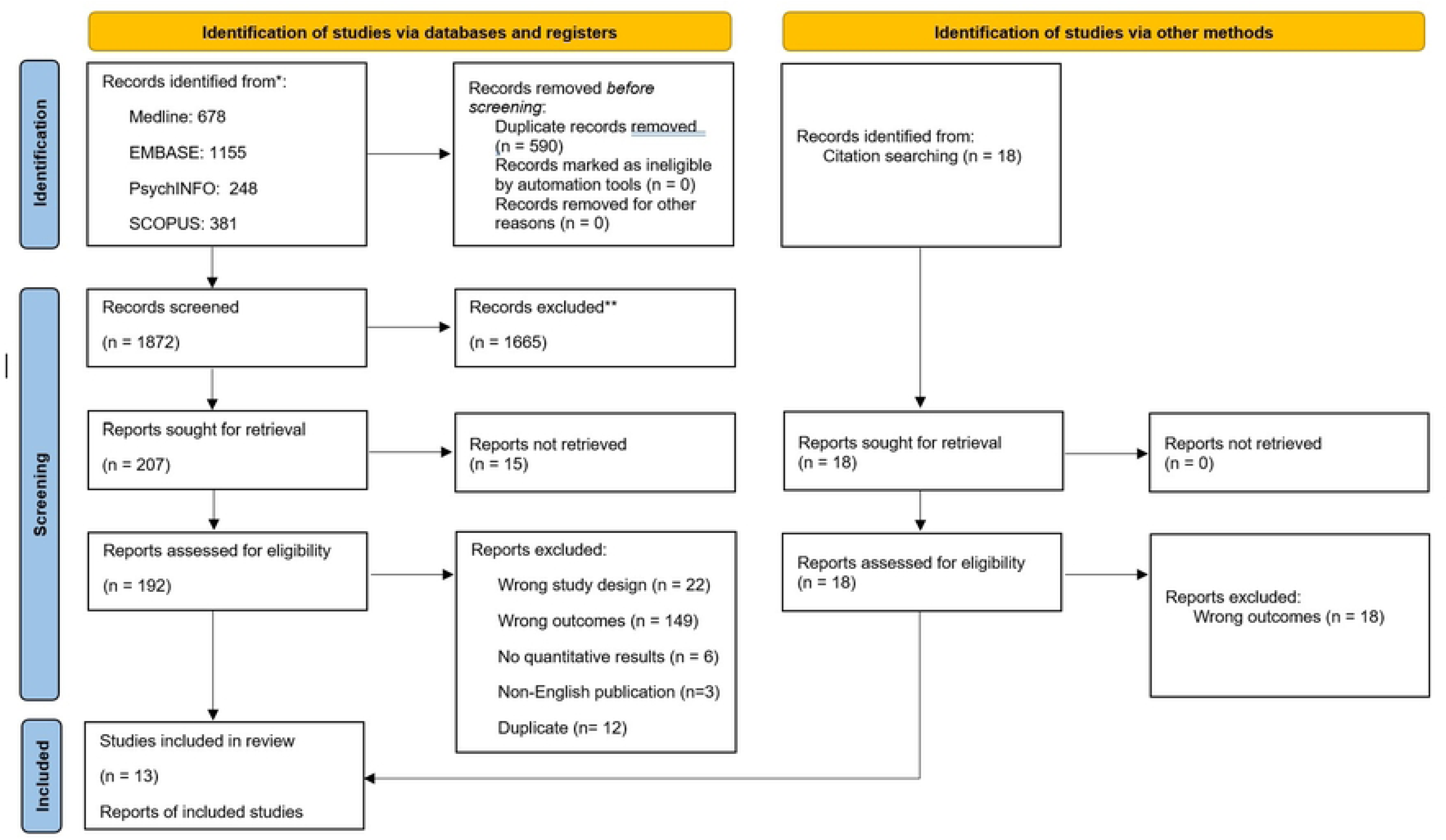
PRIMSA Flow Diagram

The characteristics of included studies are outlined in Table 1. In assessing social adversity, six studies examined low SES, three examined loneliness, two focused on psychosocial stress, and other studies examined low social network, low social capital, social isolation, and social strain. To assess cognition, six studies looked at overall cognition measured by the Digital Symbol Substitution Test (DSST), Mini-Mental State Examination (MMSE), or telephone interview for cognitive status, five studies investigated specific domains of cognition including verbal fluency, attention, memory, motor skills, executive function, and processing speed. Two studies looked at cortical morphologies and white matter infrastructure using magnetic resonance imaging. Biological mechanisms included assessments of allostatic load, inflammation, and genetics or epigenetics or genetic aging markers. Overall findings from each of the studies included are summarized in Table 2. Below, we provide a narrative summarizing the biological mechanisms linking social adversity to cognitive outcomes based on the findings of the studies included in this review.

**Table 1.**
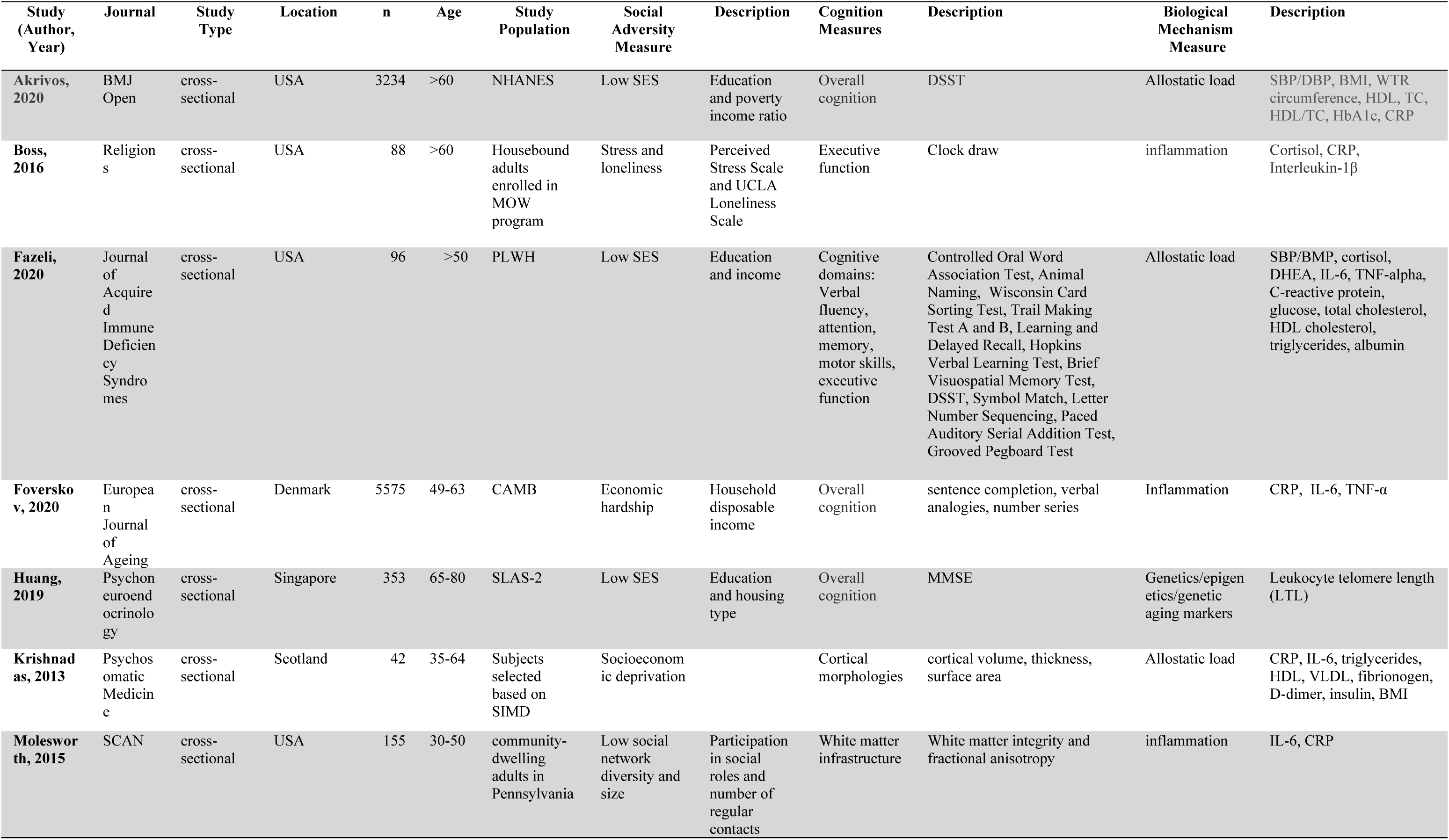
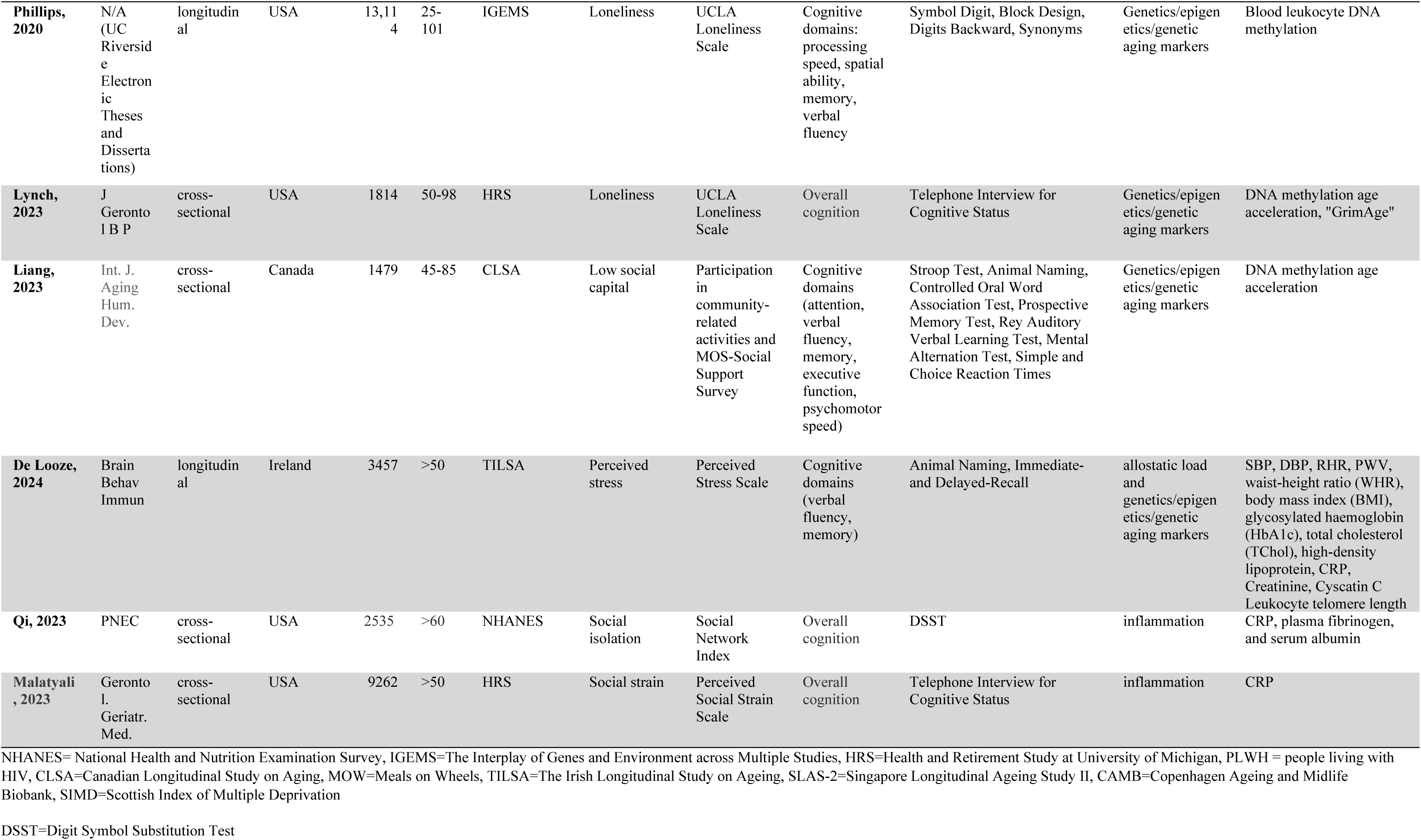
Characteristics of included studies.

**Table 2.** Findings from included studies.

| Study (Author, Year) | Association between Cognition and Social Adversity | Association between Biological Mechanism and Social Adversity | Association between Biological Mechanism and Cognition | Biological Mechanism as mediator for association between social factor and cognition |
| --- | --- | --- | --- | --- |
| Akrivos, 2020 | level of education and PIR was significantly positively associated with performance on the DSST | SES was significantly negatively associated with AL | AL was significantly negatively associated with cognitive performance | AL was not a significant mediator of the SES-related differences in cognition, it mediated at most 4.5% of the SES effect on DDST performance in the highest PIR quartile |
| Boss, 2016 | | Greater loneliness predicted greater CRP and had a near significant contribution to cortisol. | IL-1β showed a significant positive correlation with executive function. | |
| Fazeli, 2020 | SES was a significant predictor of cognitive functions. | No significant associations emerged between SES and AL. | No significant associations between AL and cognition in the overall sample, but higher AL was associated with lower cognitive outcomes in African American PLWH. | The indirect effect of SES on neurocognitive functioning through allostatic load was significant, suggesting mediation. |
| Foverskov, 2020 | Experiencing EH for more than four years was associated with significantly lower cognitive test scores. | Four or more years in EH was related to higher inflammatory levels for CRP and Interleukin-6. |  |  |
| Huang, 2019 | High SES appears to protect individuals from age-related declines in cognitive function. There is significant negative association between age and cognitive function in lower SES levels but not higher SES levels. | Among older adults with lower and mean SES levels, there is a significant age-related decline in LTL. In contrast, there is no significant association between age and LTL among individuals with higher SES. |  | Specifically, older adults with lower SES experience large age-related decreases in cognitive functioning, which are then associated with shorter LTL. In contrast, participants with higher SES maintain higher cognitive functioning despite increasing age, which in turn is related to greater LTL. |
| Krishnadas, 2013 | The MD (most deprived) group had statistically significantly smaller volumes pertaining to the left posterior parietal cortex and right Broca homologue compared with the LD (least deprived) group. |  |  | Inflammation factor mediated the relationship between deprivation status and left Wernicke's region CT. |
| Molesworth, 2015 | There is a positive association between a measure of white matter integrity, fractional anisotropy (FA), and social network diversity, particularly near anterior corpus callosum. | IL-6 was negatively associated with the diversity of a person's social network. | FA is weakly associated with levels of IL-6. | IL-6 did not mediate the social network and FA relationship. |
| Phillips, 2020 | Loneliness was negatively associated with spatial abilities, processing speed, verbal comprehension, and working memory. |  | No significant association between DNA methylation and spatial ability. Significant association with processing speed and verbal comprehension at specific CpG sites. | Although direct effects of loneliness on cognition were not significant, indirect associations of perceived loneliness to methylation of cg00403457 in PTPN12 to change in processing speed were observed, indicative of a potential role of methylation at this site in the loneliness—cognition relation. |
| **Lynch, 2023** | Significant effect of loneliness on overall cognition. |  | Significant correlations between DNAm AgeAccel measures and cognition. | GrimAge Accel consistently explained the association between loneliness and general cognitive ability, immediate recall, and delayed memory recall. |
| **Liang, 2023** | Lower structural social capital was significantly associated with worse attention and verbal fluency. | Lower structural social capital was significantly associated with greater epigenetic age acceleration difference. |  |  |
| **De Looze, 2024** | Higher perceived stress was significantly associated with worse cognitive outcomes. |  | Longitudinally, AL was associated with lower cognitive functions. | The strength of associations between perceived stress and cognitive outcomes did not change when AL was adjusted. |
| **Qi, 2023** | Socially isolated older adults were found to have a poorer cognitive functioning than those not isolated. | Social isolation was related to higher levels of CRP and fibrinogen in men. |  | The association between social isolation and cognitive functioning mediated by CRP and fibrinogen was 6.1% and 12.0%, respectively. |
| **Malatyali, 2023** | Higher level of social strain from friends was significantly associated with the risk of CIND (cognitive impairment without dementia) and dementia. |  | Higher levels of CRP were significantly associated with CIND, but not dementia. |  |

### 3.2 Inflammation

Five studies measured inflammation specifically and its relationship to social adversity and cognition. Qi et al. found that chronic inflammation was a mediator between social adversity and cognitive function. Specifically, they showed that socially isolated older adults have worse cognitive function measured by the DSST (β = −2.445, SE=1.180, p < 0.01 for men; β = −5.478, SE=1.167, p < 0.001 for women). For men specifically, the association between social isolation and cognitive functioning was mediated by c-reactive protein (CRP) and fibrinogen levels with the proportion mediated being 6.1% and 12.0%, respectively. On the other hand, Molesworth et al. assessed social networks and white matter microstructure, and found that, while there is an association between white matter integrity and social network diversity (mean=0.012, p<0.025), inflammatory cytokine interleukin-6 did not mediate this relationship (p>0.1). Boss et al. found that both social adversity and cognition were independently related to inflammation. They found that greater loneliness correlated with CRP (r=0.26, p=0.02 and cortisol levels (p=0.06), while executive function was correlated with interleukin-1β (r = 0.23, p = 0.03). Foverskov et al. similarly found that four or more years of economic hardship was related to higher levels of inflammatory biomarkers: 22% for CRP (95% CI [4, 44]) and 23% for interleukin-6 (95% CI [10., 39]). Malatyali et al. found that CRP was related to mild cognitive impairment (β = 1.03, 95% CI [1.01, 1.06], p<0.01) but not dementia (β=1.021, 95% CI [0.98, 1.97]).

### 3.3 Allostatic load

Four studies examined various markers of allostatic load as it relates to social adversity and cognition. Fazeli et al. specifically examined older adults living with HIV and found SES was a significant predictor of neurocognitive functioning (β=1.12, SE=0.41, p=0.008), and that allostatic load was a significant mediator of this relationship (β=0.19, SE=0.12, 95% CI [0.002, 0.485]). Similarly, Krishnadas et al. examined socioeconomic deprivation and measured cortical morphologies to find morphological differences between most-deprived and least-deprived groups, including a thinner left Wernicke’s area in the most-deprived (Cohen’s d=0.93), and found inflammatory markers in particular mediated this correlation (fibrinogen, interleukin-6, CRP, and D-dimers). No other cardiometabolic factors mediated the relationship between deprivation status and Wernicke’s area cortical thickness (β = −0.029, SE = 0.15, 95% CI [−0.06 to −0.007]), which is the brain region responsible for language comprehension. Meanwhile, Akrivos et al. found that poverty and education were social factors that were significantly associated with outcomes on the Digit Symbol Substitution Test (β=8.9, 95% CI [6.6 to 11.3], p<0.0001), but that allostatic load was not a significant mediator of SES-related differences in cognition because it mediated at most 4.5% of the effects and only in the highest poverty income ratio quartile. Similarly, De Looze et al. found that perceived stress was significantly associated with cognition in the domain of verbal fluency and memory (β = −0.10, 95 % CI [−0.12; −0.07], p <.001), but that the strength of these associations did not change when allostatic load was adjusted for (p=0.13).

### 3.4 Genetics, epigenetics, and genetic aging markers

Five studies examined different genetics and epigenetic markers as related to social adversity and cognition. Huang et al. and De Looze et al. both looked at leukocyte telomere length (LTL). Huang et al. found that for adults with lower SES levels, there was a negative association between age and cognitive function as measured by MMSE scores (β = 0.0339, SE = 0.0123, t = 2.75, p = 0.006). LTL served as a mediator specifically in adults with low SES, where age-related decreases in cognitive functions are associated with shorter LTL. On the other hand, De Looze et al., while also finding significant effect of perceived stress on cognitive function, did not find LTL to change the strength of these associations (*X^2^*(32) = 1.6, p =.89). Phillips examined blood leukocyte DNA methylation and found it to play a mediating role between perceived loneliness and processing speed (β = -1.44, p = 0.0040). Lynch et al. and Liang et al. both looked at DNA methylation age acceleration. Lynch et al. found that loneliness negatively impacts cognition. Specifically, GrimAge acceleration consistently mediated the relationship between loneliness and general cognitive ability, immediate recall, and delayed memory recall (β = -0.19). Similarly, Liang et al. found that epigenetic aging measured using Hannum clock in a large, population-based Canadian sample was independently associated with low structural cognitive capital (β = -0.79, 95% CI [-1.5, -0.19], p <0.05).

## 4. Discussion

This scoping review sought to investigate the biological mechanisms through which social adversity may impact cognitive health during aging. Collectively, the findings indicate that various forms of social adversity—including low socioeconomic status (SES), loneliness, and psychosocial stress—are associated with negative cognitive outcomes, particularly in older adults. Importantly, biological mechanisms such as inflammation, allostatic load, and genetic/epigenetic markers were identified as potential mediators in these relationships, though the strength and consistency of these associations varied across studies. Inflammation appeared to play a prominent role, with several studies demonstrating that higher levels of inflammatory markers, such as CRP and interleukin-6, were linked to both social adversity and cognitive decline(32,37). However, other studies did not find inflammation to mediate the relationship between social adversity and cognition, highlighting the complexity of these interactions(35). These mixed findings suggest that while inflammation is a plausible biological pathway, it may not be universally relevant for all individuals or social contexts, and further research is needed to better delineate its role in cognitive aging.

Allostatic load, a measure of the cumulative physiological wear and tear resulting from chronic stress, was another biological mechanism identified in the reviewed studies. Four studies explored this concept in the context of social adversity and cognitive outcomes. For instance, research by Fazeli et al. (2021) demonstrated that allostatic load mediated the relationship between SES and neurocognitive function in older adults living with HIV, supporting the notion that prolonged exposure to social adversity may lead to physiological dysregulation, which in turn exacerbates cognitive decline(30). Similarly, Krishnadas et al. (2013) found that socioeconomic deprivation was associated with cortical structural changes, particularly in the Wernicke’s area, and that inflammatory markers mediated this relationship(34). Conversely, Akrivos et al. (2020) and De Looze et al. (2021) reported that while social adversity was associated with poorer cognitive performance, allostatic load explained only partly explained the proportion of this variance, suggesting that other mechanisms may be more central to these associations(28,39). These findings point to the need for a more nuanced understanding of how allostatic load contributes to cognitive aging, particularly when considered alongside other factors such as inflammation and genetic predispositions, and importantly social and environmental exposures in early-life and across the life course(40).

The role of genetics, epigenetics, and genetic aging markers in mediating the relationship between social adversity and cognitive outcomes emerged as another important area of interest. Several studies investigated telomere length (LTL), DNA methylation, and epigenetic aging as potential biological mediators. Huang et al. (2020) found that shorter LTL mediated the relationship between low SES and cognitive decline, providing support for the hypothesis that social adversity accelerates biological aging. However, other studies, such as De Looze et al. (2021), did not find LTL to mediate the association between perceived stress and cognitive outcomes, suggesting that epigenetic mechanisms may be context-dependent. Notably, studies examining DNA methylation, including those by Phillips et al. (2020) and Liang et al. (2021), found that specific epigenetic changes, such as those linked to loneliness and social isolation, were associated with poorer cognitive function. For example, Phillips et al. (2020) showed that DNA methylation in blood leukocytes mediated the effect of loneliness on processing speed, while Lynch et al. (2020) demonstrated that epigenetic aging, as measured by GrimAge, mediated the relationship between loneliness and cognitive decline. These findings suggest that epigenetic alterations could represent a critical biological pathway through which social adversity influences cognitive health, though more research is needed to establish causal links and to identify which epigenetic markers are most predictive of cognitive outcomes.

Notably, while inflammation, allostatic load, and epigenetics represent promising biological pathways linking social adversity to cognitive decline, the reviewed studies also highlight the need for more comprehensive models that integrate multiple biological, psychological, and social factors. Many of the studies included in this review focused on a single biological mechanism or social factor, often in isolation, and did not consider the potential for complex interactions between these variables. For instance, while inflammation was a common mediator in the studies examining social isolation, its effect was not always consistent across different cognitive domains or social contexts(32,37). Similarly, while allostatic load and SES were found to be associated with cognitive outcomes, the magnitude of the relationship was often modest and varied depending on the individual’s specific life circumstances(28). This suggests that future research should adopt a more integrative, multidimensional approach that considers the dynamic interplay between genetic, physiological, and social factors in shaping cognitive health(41). Longitudinal studies with larger and more diverse populations are particularly needed to establish causal relationships and to explore how these pathways evolve over time, especially in light of the aging process. By advancing our understanding of the complex mechanisms through which social adversity affects cognitive aging, researchers can help inform interventions aimed at mitigating the impact of social inequalities on cognitive health in older adults.

There are several strengths to our study. Our search strategy was developed in collaboration with a trained research librarian to identify the optimal data sources and search terms for a comprehensive review. Quality assessment showed that all the studies included in our review achieved over seven points on the nine-point NOS scale. This is largely since most studies employed pre-existing, large-scale, nationally representative data, minimizing selection bias in participants and confirmation bias in data analysis. Our study also has some limitations including the modest number of studies that were eligible for inclusion. Also, the heterogeneity in the studies included in terms of social adversity, cognitive domains, and biological factors may have limited the ability to compare the included studies. Our review also excluded studies in languages other than English limiting global representation.

### Clinical and health policy implications

The findings of this scoping review have some clinical and health policy implications, particularly for addressing cognitive decline and dementia in aging populations. Clinically, the recognition of social adversity—such as low SES, social isolation, and chronic stress—as significant risk factors for cognitive decline highlights the need for healthcare providers to take a holistic, biopsychosocial approach when assessing and managing older adults. Incorporating social assessments into routine cognitive screenings could help identify at-risk individuals early, allowing for timely interventions aimed at mitigating the effects of social adversity on brain health.

Additionally, given the role of biological mediators such as inflammation and allostatic load, clinicians should consider implementing strategies to reduce inflammation and managing stress in older adults, including lifestyle interventions like physical activity, social engagement, and stress management techniques, which have been shown to improve both physical and cognitive outcomes(5,42). From a policy perspective, these findings emphasize the need for public health initiatives that address the social determinants of health, particularly in vulnerable aging populations. Policies that promote social connectedness, economic stability, and access to mental health resources could help reduce the burden of cognitive decline and dementia, particularly in communities facing higher levels of socioeconomic deprivation. Moreover, policies aimed at reducing health disparities by providing equitable access to healthcare and social services for older adults are crucial in mitigating the cognitive impacts of social adversity. The integration of social interventions, such as community-based programs that foster social networks and reduce isolation, could complement traditional medical approaches to aging and cognitive health. In summary, a multidisciplinary approach—incorporating both social and biomedical interventions—should be prioritized in clinical practice and health policy to optimize cognitive aging and improve the quality of life for older adults.

## Conclusion

In conclusion, this scoping review underscores the complex and multifactorial nature of the relationship between social adversity and cognitive health in aging, highlighting the role of biological mediators such as inflammation, allostatic load, and epigenetic changes. While substantial evidence supports the influence of social factors—such as SES, loneliness, and psychosocial stress—on cognitive decline, the specific biological pathways that mediate these associations remain only partially understood. Our review reveals both the promise and the limitations of current research, pointing to the need for more comprehensive, longitudinal studies that integrate biological, psychological, and social perspectives to fully capture the dynamic interactions between these factors. By addressing the existing knowledge gaps, future research can better inform interventions and public health strategies aimed at promoting cognitive resilience and reducing the burden of cognitive decline among aging populations, particularly in socially disadvantaged groups. Ultimately, a deeper understanding of these pathways will be critical for developing targeted interventions that foster healthy aging and improve cognitive outcomes for individuals across the life course.

## Data Availability

All relevant data are within the manuscript and its Supporting Information files.

## CReDiT statement

Aileen Liang: Conceptualization, Methodology, Formal Analysis, Investigation, Data Curation, Writing - Original Draft, Writing - Review & Editing, Visualization

Grace Watt: Methodology, Formal Analysis, Data Curation

Noha Gomaa: Conceptualization, Methodology, Writing - Original Draft, Writing - Review & Editing, VIsualization, Supervision, Funding Acquisition

## Acknowledgements

AL was supported by the Summer Research Training Program (SRTP) at Western University, Canada, during the completion of this study. NG is supported by funding from the Canadian Institutes of Health Research (CIHR) and the Schulich School of Medicine & Dentistry, Western University

## S1 Appendix. Complete Search Strategy

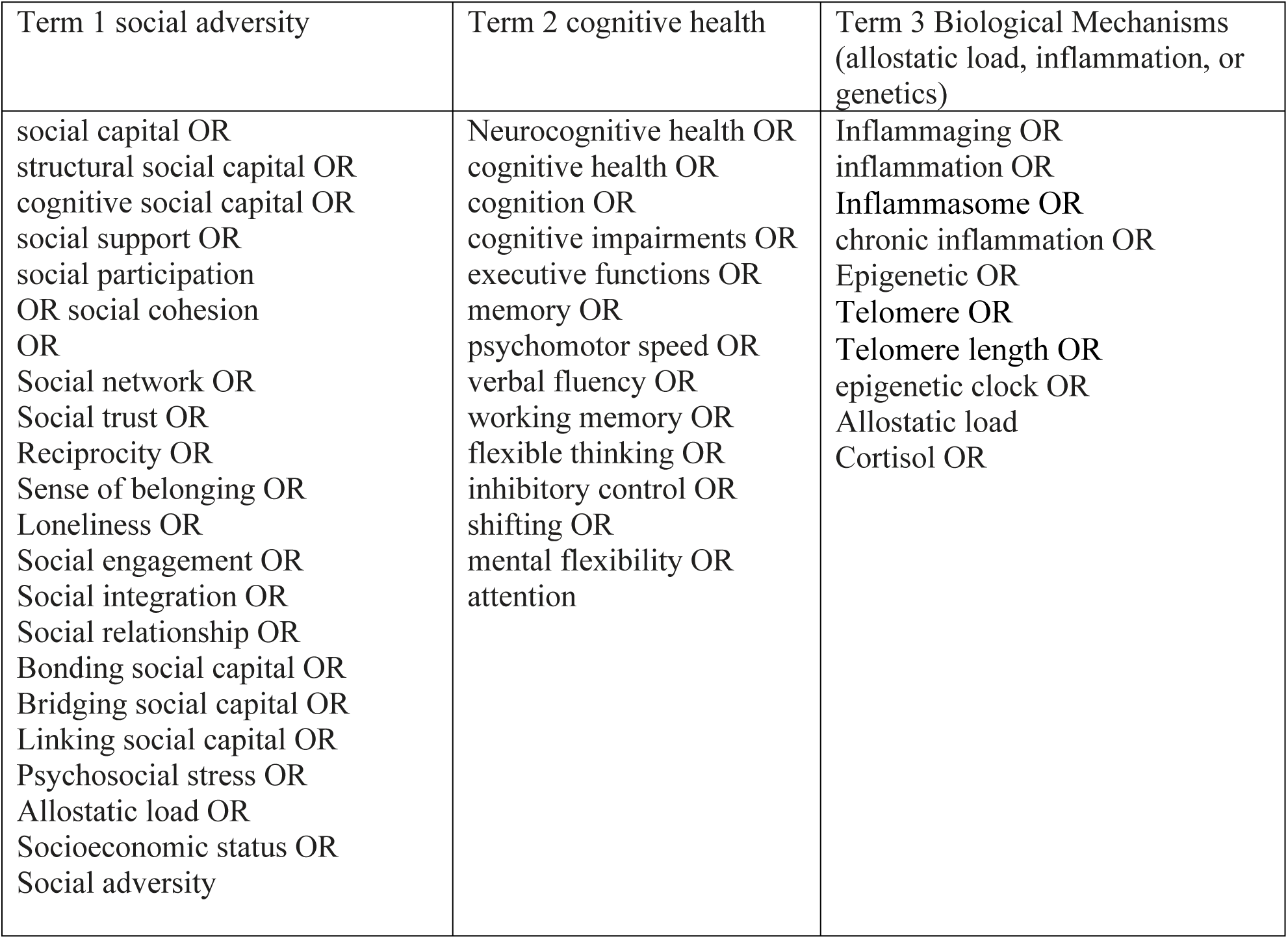

1722 articles imported, 590 duplicates removed.

### Databases

- Medline (from Ovid) - 616
- Embase (from Ovid) - 1080
- psychINFO (from Ovid) - 248
- SCOPUS – 368

References of review articles and studies were also manually searched for additional studies.

**Cutoff date: March 5, 2024**

### Total

**Total:**
| **Level 1 screening (title and abstract)** |
| --- |
| • **Age: over 18**<br>• **Country: any**<br>• **Language: English/translated**<br>• **Must contain a measure of cognition or a measure of inflammaging**<br>• **Must contain a measure of social adversity** |

| **Inclusion criteria** | **Exclusion criteria** |
| --- | --- |
| • **Age: over 18**<br>• **Country: any**<br>• **Language: English**<br>• **Study type: any primary research**<br>• **Must contain a measure of cognition or a measure of biological mechanism**<br>• **Must contain a measure of social adversity (low social capital, low SES, psychosocial stress)** | • **Reviews/meta-analysis**<br>• **Studies focused on pediatric population or early childhood events/exposures**<br>• **Studies in languages other than English**<br>• **Mentions of stress that does not pertain to psychosocial environment (eg. Acute, lab-induced stress or psychiatric/mood disorders)**<br>• **Non-human study** |

Medline (from Ovid)

- Social capital + cognitive health + epigenetics: 70
- Other social factors + cognitive health + epigenetics: 514

Embase

- Social capital + cognitive health + epigenetics: 433
- Other social factors + cognitive health + epigenetics: 641

PsychINFO

- Social capital + cognitive health + epigenetics: 97
- Other social factors + cognitive health + epigenetics: 359

Scopus

- Social capital + cognitive health + epigenetics: 187
- Other social factors + cognitive health + epigenetics: 161

### Medline

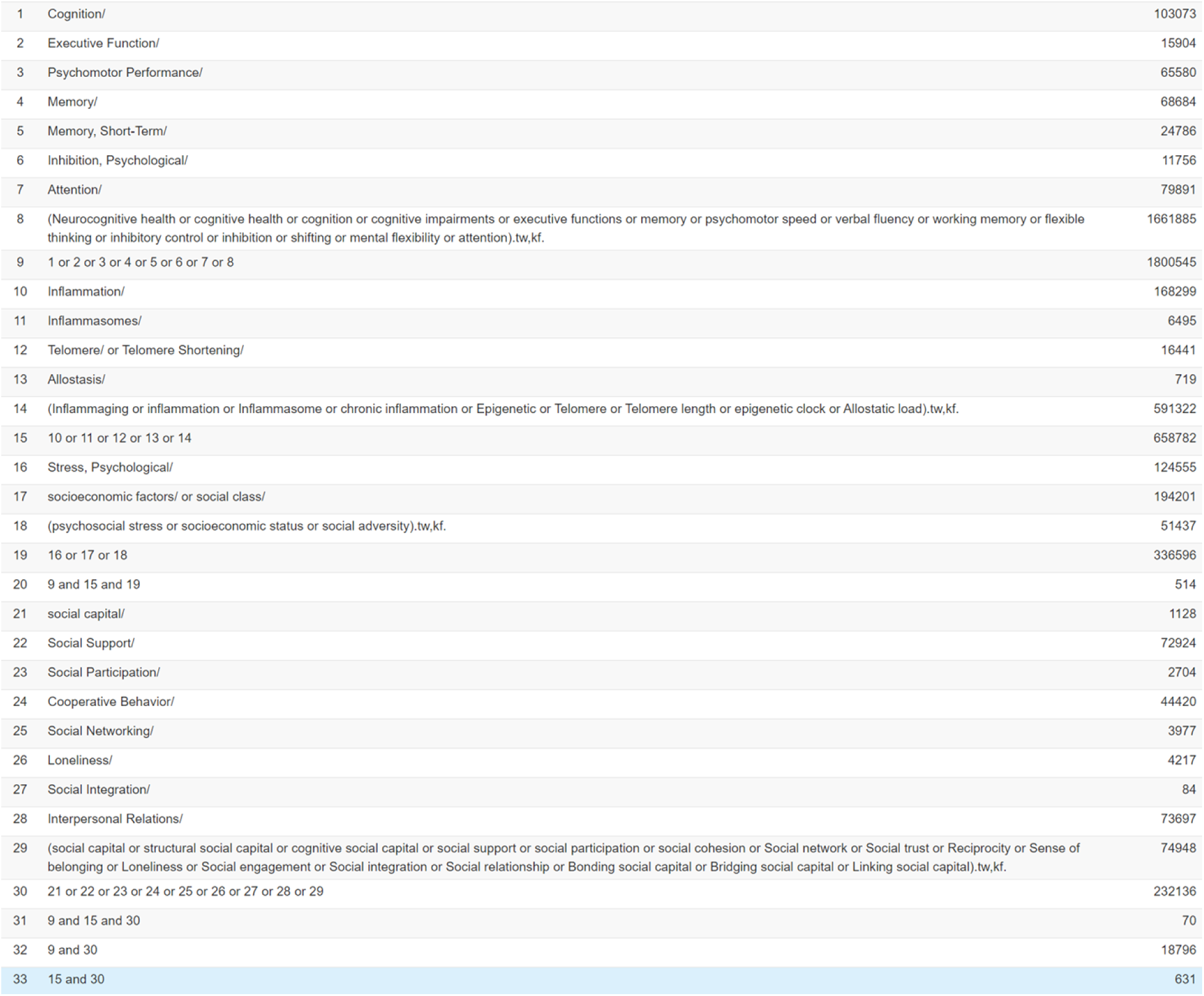

|  |  |  |
| --- | --- | --- |
| 1 | Cognition/ | 103073 |
| 2 | Executive Function/ | 15904 |
| 3 | Psychomotor Performance/ | 65580 |
| 4 | Memory/ | 68684 |
| 5 | Memory, Short-Term/ | 24786 |
| 6 | Inhibition, Psychological/ | 11756 |
| 7 | Attention/ | 79891 |
| 8 | (Neurocognitive health or cognitive health or cognition or cognitive impairments or executive functions or memory or psychomotor speed or verbal fluency or working memory or flexible thinking or inhibitory control or inhibition or shifting or mental flexibility or attention).tw,kf. | 1661885 |
| 9 | 1 or 2 or 3 or 4 or 5 or 6 or 7 or 8 | 1800545 |
| 10 | Inflammation/ | 168299 |
| 11 | Inflammasomes/ | 6495 |
| 12 | Telomere/ or Telomere Shortening/ | 16441 |
| 13 | Allostasis/ | 719 |
| 14 | (Inflammaging or inflammation or Inflammasome or chronic inflammation or Epigenetic or Telomere or Telomere length or epigenetic clock or Allostatic load).tw,kf. | 591322 |
| 15 | 10 or 11 or 12 or 13 or 14 | 658782 |
| 16 | Stress, Psychological/ | 124555 |
| 17 | socioeconomic factors/ or social class/ | 194201 |
| 18 | (psychosocial stress or socioeconomic status or social adversity).tw,kf. | 51437 |
| 19 | 16 or 17 or 18 | 336596 |
| 20 | 9 and 15 and 19 | 514 |
| 21 | social capital/ | 1128 |
| 22 | Social Support/ | 72924 |
| 23 | Social Participation/ | 2704 |
| 24 | Cooperative Behavior/ | 44420 |
| 25 | Social Networking/ | 3977 |
| 26 | Loneliness/ | 4217 |
| 27 | Social Integration/ | 84 |
| 28 | Interpersonal Relations/ | 73697 |
| 29 | (social capital or structural social capital or cognitive social capital or social support or social participation or social cohesion or Social network or Social trust or Reciprocity or Sense of belonging or Loneliness or Social engagement or Social integration or Social relationship or Bonding social capital or Bridging social capital or Linking social capital).tw,kf. | 74948 |
| 30 | 21 or 22 or 23 or 24 or 25 or 26 or 27 or 28 or 29 | 232136 |
| 31 | 9 and 15 and 30 | 70 |
| 32 | 9 and 30 | 18796 |
| 33 | 15 and 30 | 631 |

### EMBASE

| ☐ | # ▲ | Searches | Results | Type |
| --- | --- | --- | --- | --- |
| ☐ | 1 | social capital/ | 3129 | Advanced |
| ☐ | 2 | social support/ | 96427 | Advanced |
| ☐ | 3 | social participation/ | 6731 | Advanced |
| ☐ | 4 | social network/ | 18486 | Advanced |
| ☐ | 5 | trust/ | 26925 | Advanced |
| ☐ | 6 | loneliness/ | 9831 | Advanced |
| ☐ | 7 | social interaction/ | 58641 | Advanced |
| ☐ | 8 | (social capital or structural social capital or cognitive social capital or social support or social participation or social cohesion or Social network or Social trust or Reciprocity or Sense of belonging or Loneliness or Social engagement or Social integration or Social relationship or Bonding social capital or Bridging social capital or Linking social capital).tw,kw. | 93895 | Advanced |
| ☐ | 9 | 1 or 2 or 3 or 4 or 5 or 6 or 7 or 8 | 235895 | Advanced |
| ☐ | 10 | cognition/ | 244844 | Advanced |
| ☐ | 11 | cognitive defect/ | 175558 | Advanced |
| ☐ | 12 | executive function/ | 43589 | Advanced |
| ☐ | 13 | memory/ | 147449 | Advanced |
| ☐ | 14 | psychomotor performance/ | 23991 | Advanced |
| ☐ | 15 | (Neurocognitive health or cognitive health or cognition or cognitive impairments or executive functions or memory or psychomotor speed or verbal fluency or working memory or flexible thinking or inhibitory control or inhibition or shifting or mental flexibility or attention).tw,kw. | 2210138 | Advanced |
| ☐ | 16 | inflammation/ | 497202 | Advanced |
| ☐ | 17 | inflammasome/ | 16532 | Advanced |
| ☐ | 18 | chronic inflammation/ | 37207 | Advanced |
| ☐ | 19 | epigenetics/ | 85087 | Advanced |
| ☐ | 20 | allostasis/ | 881 | Advanced |
| ☐ | 21 | (Inflammaging or inflammation or Inflammasome or chronic inflammation or Epigenetic or Telomere or Telomere length or epigenetic clock or Allostatic load).tw,kw. | 892944 | Advanced |
| ☐ | 22 | 16 or 17 or 18 or 19 or 20 or 21 | 1102956 | Advanced |
| ☐ | 23 | 10 or 11 or 12 or 13 or 14 or 15 | 2476203 | Advanced |
| ☐ | 24 | 9 and 22 and 23 | 403 | Advanced |
| ☐ | 25 | 9 and 22 | 1849 | Advanced |
| ☐ | 26 | 9 and 23 | 30368 | Advanced |
| ☐ | 27 | mental stress/ | 90240 | Advanced |
| ☐ | 28 | social status/ | 100485 | Advanced |
| ☐ | 29 | (psychosocial stress or socioeconomic status or social adversity).tw,kw. | 61639 | Advanced |
| ☐ | 30 | 27 or 28 or 29 | 215270 | Advanced |
| ☐ | 31 | 22 and 23 and 30 | 613 | Advanced |

### PsychINFO

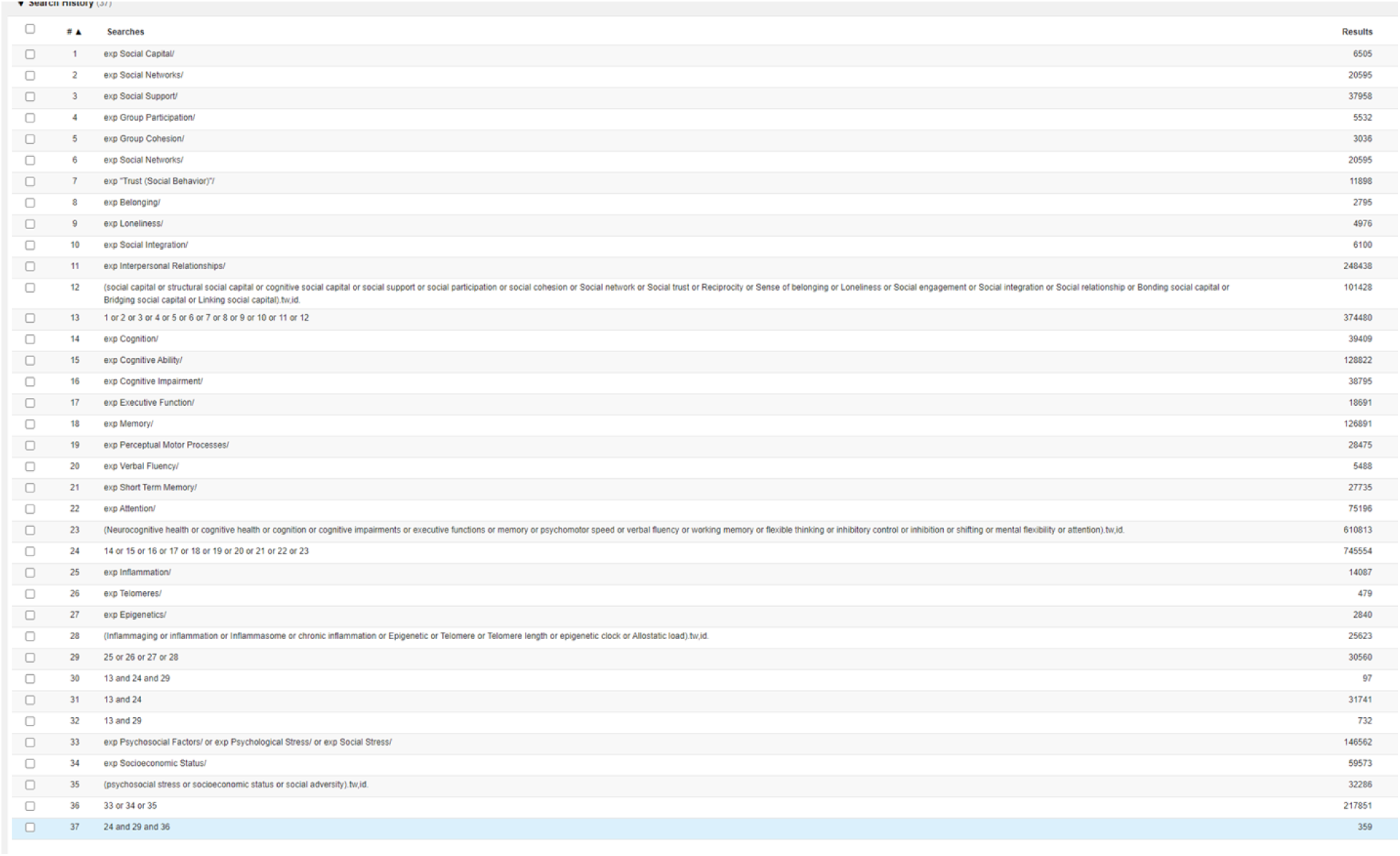

### Scopus

(TITLE-ABS-KEY ("social capital" OR "structural social capital" OR "cognitive social capital" OR "social support" OR "social participation" OR "social cohesion" OR "Social network" OR "Social trust" OR "Reciprocity" OR "Sense of belonging" OR "Loneliness" OR "Social engagement" OR "Social integration" OR "Social relationship" OR "Bonding social capital" OR "Bridging social capital" OR "linking social capital") AND TITLE-ABS-KEY ("Neurocognitive health" OR "cognitive health" OR "cognition" OR "cognitive impairment*" OR "executive function*" OR "memory" OR "psychomotor speed" OR "verbal fluency" OR "working memory" OR "flexible thinking" OR "inhibitory control" OR "inhibition" OR "shifting" OR "mental flexibility" OR "attention"))

(TITLE-ABS-KEY ("social capital" OR "structural social capital" OR "cognitive social capital" OR "social support" OR "social participation" OR "social cohesion" OR "Social network" OR "Social trust" OR "Reciprocity" OR "Sense of belonging" OR "Loneliness" OR "Social engagement" OR "Social integration" OR "Social relationship" OR "Bonding social capital" OR "Bridging social capital" OR "linking social capital") AND TITLE-ABS-KEY ("Neurocognitive health" OR "cognitive health" OR "cognition" OR "cognitive impairment*" OR "executive function*" OR "memory" OR "psychomotor speed" OR "verbal fluency" OR "working memory" OR "flexible thinking" OR "inhibitory control" OR "inhibition" OR "shifting" OR "mental flexibility" OR "attention") AND TITLE-ABS-KEY ("Inflammaging" OR "inflammation" OR "Inflammasome" OR "chronic inflammation" OR "Epigenetic*" OR "Telomere*" OR "Telomere length" OR "epigenetic clock" OR "Allostatic load"))

(TITLE-ABS-KEY ("Inflammaging" OR "inflammation" OR "Inflammasome" OR "chronic inflammation" OR "Epigenetic*" OR "Telomere*" OR "Telomere length" OR "epigenetic clock" OR "Allostatic load") AND TITLE-ABS-KEY ("Neurocognitive health" OR "cognitive health" OR "cognition" OR "cognitive impairment*" OR "executive function*" OR "memory" OR "psychomotor speed" OR "verbal fluency" OR "working memory" OR "flexible thinking" OR "inhibitory control" OR "inhibition" OR "shifting" OR "mental flexibility" OR "attention") AND TITLE-ABS-KEY ("psychosocial stress" OR "socioeconomic status" OR "social adversit*"))

## S2 Appendix. Results of Newcastle-Ottawa Quality Assessment Scale

|  | Selection |  |  |  | Comparability |  | Outcome |  |  |
| --- | --- | --- | --- | --- | --- | --- | --- | --- | --- |
|  | Representativeness of the exposed cohort | Selection of the non exposed cohort | Ascertainment of exposure | Demonstration that outcome of interest was not present at start of study | Study controls for age and sex | Study controls for additional factors | Assessment of outcome | Was follow-up long enough for outcomes to occur | Adequacy of follow up of cohorts |
| Akrivos 2020 | * | * | * | * | * | * | * | * | * |
| Boss 2016 | * | * | * | 0 | * | * | * | * | * |
| Fazeli 2020 | * | * | 0 | * | * | * | * | * | * |
| Foverskov 2020 | * | * | * | * | * | * | * | * | * |
| Hatton 2018 | * | * | * | 0 | * | * | * | * | * |
| Huang 2019 | * | * | 0 | * | * | 0 | * | * | * |
| Krishnadas 2013 | * | * | * | 0 | * | 0 | * | * | * |
| Wilson 2007 | * | * | * | * | * | * | * | * | * |
| Molesworth 2015 | * | 0 | 0 | * | * | * | * | * | * |
| Phillips 2020 | * | 0 | * | * | * | * | * | * | * |
| Lynch 2023 | * | * | * | * | * | * | * | * | * |
| Liang 2023 | * | * | * | * | * | * | * | * | * |
| De Looze 2024 | * | * | * | * | * | * | * | * | * |
| Qi 2023 | * | * | * | * | * | * | * | * | * |
| Malatyali 2023 | * | * | * | * | * | * | * | * | * |

## References

1. Kelly ME, Duff H, Kelly S, McHugh Power JE, Brennan S, Lawlor BA, et al. The impact of social activities, social networks, social support and social relationships on the cognitive functioning of healthy older adults: a systematic review. Syst Rev [Internet]. 2017 Dec 19 [cited 2025 Mar 20];6(1). Available from: https://pubmed.ncbi.nlm.nih.gov/29258596/

2. Seeman TE, Lusignolo TM, Albert M, Berkman L. Social relationships, social support, and patterns of cognitive aging in healthy, high-functioning older adults: MacArthur studies of successful aging. Health Psychol [Internet]. 2001 [cited 2025 Mar 20];20(4):243–55. Available from: https://pubmed.ncbi.nlm.nih.gov/11515736/

3. James BD, Wilson RS, Barnes LL, Bennett DA. Late-life social activity and cognitive decline in old age. J Int Neuropsychol Soc [Internet]. 2011 Nov [cited 2025 Mar 20];17(6):998–1005. Available from: https://pubmed.ncbi.nlm.nih.gov/22040898/

4. Cacioppo JT, Cacioppo S. Social Relationships and Health: The Toxic Effects of Perceived Social Isolation. Soc Personal Psychol Compass [Internet]. 2014 Feb 1 [cited 2025 Mar 20];8(2):58–72. Available from: https://onlinelibrary.wiley.com/doi/full/10.1111/spc3.12087

5. Wilson RS, Scherr PA, Schneider JA, Tang Y, Bennett DA. Relation of cognitive activity to risk of developing Alzheimer disease. Neurology [Internet]. 2007 [cited 2025 Mar 22];69(20):1911–20. Available from: https://pubmed.ncbi.nlm.nih.gov/17596582/

6. Hughes TF, Flatt JD, Fu B, Chang CCH, Ganguli M. Engagement in Social Activities and Progression from Mild to Severe Cognitive Impairment: The MYHAT Study. Int Psychogeriatr [Internet]. 2012 Apr [cited 2025 Mar 22];25(4):587. Available from: https://pmc.ncbi.nlm.nih.gov/articles/PMC3578022/

7. Hallman S, LeVasseur S, Bérard-Chagnon J, Martel L. A portrait of Canada’s growing population aged 85 and older from the 2021 Census [Internet]. Statistics Canada. 2022 [cited 2022 Jul 5]. Available from: https://www12.statcan.gc.ca/census-recensement/2021/as-sa/98-200-X/2021004/98-200-X2021004-eng.cfm

8. Andrianopoulos V, Gloeckl R, Vogiatzis I, Kenn K. Cognitive impairment in COPD: should cognitive evaluation be part of respiratory assessment? Breathe (Sheffield, England) [Internet]. 2017 Mar 1 [cited 2022 Jul 5];13(1):e1–9. Available from: https://pubmed-ncbi-nlm-nih-gov.proxy1.lib.uwo.ca/29184593/

9. Lara E, Caballero FF, Rico-Uribe LA, Olaya B, Haro JM, Ayuso-Mateos JL, et al. Are loneliness and social isolation associated with cognitive decline? Int J Geriatr Psychiatry [Internet]. 2019 Nov 1 [cited 2022 Jul 5];34(11):1613–22. Available from: https://pubmed-ncbi-nlm-nih-gov.proxy1.lib.uwo.ca/31304639/

10. Hackman DA, Farah MJ, Meaney MJ. Socioeconomic status and the brain: Mechanistic insights from human and animal research. Nat Rev Neurosci [Internet]. 2010 Sep [cited 2025 Mar 20];11(9):651–9. Available from: https://pubmed.ncbi.nlm.nih.gov/20725096/

11. Engel GL. The need for a new medical model: a challenge for biomedicine. Science [Internet]. 1977 [cited 2025 Mar 20];196(4286):129–36. Available from: https://pubmed.ncbi.nlm.nih.gov/847460/

12. Hensel ALJ, Nicholson K, Anderson KK, Gomaa NA. Biopsychosocial factors in oral and systemic diseases: a scoping review. Front Oral Heal. 2024 May 30;5:1378467.

13. Liang A, Gomaa N. Social Capital Associates With Better Cognitive Health, Oral Health and Epigenetic Age Deceleration: Findings From the Canadian Longitudinal Study on Aging. Int J Aging Hum Dev [Internet]. 2024 Oct 1 [cited 2025 Mar 22];99(3). Available from: https://pubmed.ncbi.nlm.nih.gov/37974418/

14. McEwen BS. Protective and damaging effects of stress mediators: central role of the brain. Dialogues Clin Neurosci [Internet]. 2006 [cited 2025 Mar 20];8(4):367. Available from: https://pmc.ncbi.nlm.nih.gov/articles/PMC3181832/

15. Hensel ALJ, Gomaa N. Social and economic capital as effect modifiers of the association between psychosocial stress and oral health. PLoS One [Internet]. 2023 May 1 [cited 2025 Mar 20];18(5):e0286006. Available from: https://journals.plos.org/plosone/article?id=10.1371/journal.pone.0286006

16. Muscatell KA, Dedovic K, Slavich GM, Jarcho MR, Breen EC, Bower JE, et al. Neural mechanisms linking social status and inflammatory responses to social stress. Soc Cogn Affect Neurosci [Internet]. 2016 Jun 1 [cited 2025 Mar 20];11(6):915–22. Available from: https://pubmed.ncbi.nlm.nih.gov/26979965/

17. Steptoe A, Hamer M, Chida Y. The effects of acute psychological stress on circulating inflammatory factors in humans: a review and meta-analysis. Brain Behav Immun [Internet]. 2007 Oct [cited 2025 Mar 22];21(7):901–12. Available from: https://pubmed.ncbi.nlm.nih.gov/17475444/

18. Calsolaro V, Edison P. Neuroinflammation in Alzheimer’s disease: Current evidence and future directions. Alzheimer’s Dement [Internet]. 2016 Jun 1 [cited 2025 Mar 22];12(6):719–32. Available from: https://onlinelibrary.wiley.com/doi/full/10.1016/j.jalz.2016.02.010

19. Faraji J, Metz GAS. Harnessing BDNF Signaling to Promote Resilience in Aging. Aging Dis [Internet]. 2024 Nov 27 [cited 2025 Mar 22];0-. Available from: https://www.aginganddisease.org/EN/10.14336/AD.2024.0961

20. Fiorito G, Polidoro S, Dugué PA, Kivimaki M, Ponzi E, Matullo G, et al. Social adversity and epigenetic aging: a multi-cohort study on socioeconomic differences in peripheral blood DNA methylation. Sci Reports 2017 71 [Internet]. 2017 Nov 24 [cited 2022 Jul 5];7(1):1–12. Available from: https://www.nature.com/articles/s41598-017-16391-5

21. Fransquet PD, Wrigglesworth J, Woods RL, Ernst ME, Ryan J. The epigenetic clock as a predictor of disease and mortality risk: A systematic review and meta-analysis. Clin Epigenetics [Internet]. 2019 Apr 11 [cited 2022 Jul 5];11(1):1–17. Available from: https://clinicalepigeneticsjournal.biomedcentral.com/articles/10.1186/s13148-019-0656-7

22. Guidi J, Lucente M, Sonino N, Fava GA. Allostatic Load and Its Impact on Health: A Systematic Review. Psychother Psychosom [Internet]. 2020 Dec 15 [cited 2025 Mar 22];90(1):11–27. Available from: 10.1159/000510696

23. Sindi S, Solomon A, Kåreholt I, Hovatta I, Antikainen R, Hänninen T, et al. Telomere Length Change in a Multidomain Lifestyle Intervention to Prevent Cognitive Decline: A Randomized Clinical Trial. Journals Gerontol Ser A [Internet]. 2021 Feb 25 [cited 2025 Mar 22];76(3):491–8. Available from: 10.1093/gerona/glaa279

24. Mcewen BS, Stellar E. Stress and the Individual Mechanisms Leading to Disease. Arch Intern Med. 1993;153:2093–101.

25. Tricco AC, Lillie E, Zarin W, O’Brien KK, Colquhoun H, Levac D, et al. PRISMA extension for scoping reviews (PRISMA-ScR): Checklist and explanation. Ann Intern Med [Internet]. 2018 Oct 2 [cited 2025 Mar 22];169(7):467–73. Available from: https://www.prisma-statement.org/scoping

26. Social determinants of health [Internet]. [cited 2025 Mar 25]. Available from: https://www.who.int/health-topics/social-determinants-of-health#tab=tab_1

27. Harvey PD. Domains of cognition and their assessment. Dialogues Clin Neurosci [Internet]. 2019 [cited 2025 Apr 3];21(3):227–37. Available from: https://pubmed-ncbi-nlm-nih-gov.proxy1.lib.uwo.ca/31749647/

28. Akrivos J, Zhu CW, Haroutunian V. Role of cumulative biological risk in mediating socioeconomic disparities in cognitive function in the elderly: a mediation analysis. BMJ Open [Internet]. 2020 Sep 18 [cited 2025 Mar 27];10(9):e035847. Available from: https://pmc.ncbi.nlm.nih.gov/articles/PMC7511641/

29. Lynch M, Em Arpawong T, Beam CR. Associations Between Longitudinal Loneliness, DNA Methylation Age Acceleration, and Cognitive Functioning. J Gerontol B Psychol Sci Soc Sci [Internet]. 2023 Dec 1 [cited 2025 Mar 28];78(12):2045–59. Available from: https://pubmed.ncbi.nlm.nih.gov/37718577/

30. Fazeli PL, Waldrop-Valverde D, Yigit I, Turan B, Edberg J, Kempf M, et al. An exploratory study of correlates of allostatic load in older people living with HIV. J Acquir Immune Defic Syndr [Internet]. 2020 Apr 15 [cited 2025 Mar 28];83(5):441. Available from: https://pmc.ncbi.nlm.nih.gov/articles/PMC7424692/

31. Foverskov E, Petersen GL, Pedersen JLM, Rod NH, Mortensen EL, Bruunsgaard H, et al. Economic hardship over twenty-two consecutive years of adult life and markers of early ageing: physical capability, cognitive function and inflammation. Eur J Ageing [Internet]. 2020 Mar 1 [cited 2025 Mar 27];17(1):55–67. Available from: https://link-springer-com.proxy1.lib.uwo.ca/article/10.1007/s10433-019-00523-z

32. Boss L, Branson S, Cron S, Kang DH. Biobehavioral Examination of Religious Coping, Psychosocial Factors, and Executive Function in Homebound Older Adults. Relig 2016, Vol 7, Page 42 [Internet]. 2016 Apr 26 [cited 2025 Mar 27];7(5):42. Available from: https://www.mdpi.com/2077-1444/7/5/42/htm

33. Huang Y, Yim OS, Lai PS, Yu R, Chew SH, Gwee X, et al. Successful aging, cognitive function, socioeconomic status, and leukocyte telomere length. Psychoneuroendocrinology [Internet]. 2019 May 1 [cited 2025 Mar 27];103:180–7. Available from: https://pubmed-ncbi-nlm-nih-gov.proxy1.lib.uwo.ca/30708136/

34. Krishnadas R, Mclean J, Batty GD, Burns H, Deans KA, Ford I, et al. Socioeconomic deprivation and cortical morphology: psychological, social, and biological determinants of ill health study. Psychosom Med [Internet]. 2013 [cited 2025 Mar 27];75(7):616–23. Available from: https://pubmed.ncbi.nlm.nih.gov/23975946/

35. Molesworth T, Sheu LK, Cohen S, Gianaros PJ, Verstynen TD. Social network diversity and white matter microstructural integrity in humans. [cited 2025 Mar 27]; Available from: http://www.psy.cmu.

36. Phillips DM. UC Riverside UC Riverside Electronic Theses and Dissertations Title Longitudinal Loneliness and Cognitive Aging in Mid and Late Life: Patterns of Associations and Epigenetic Pathways [Internet]. 2020 [cited 2025 Mar 27]. Available from: https://escholarship.org/uc/item/7891d9wn

37. Qi X, Ng TKS, Wu B. Sex differences in the mediating role of chronic inflammation on the association between social isolation and cognitive functioning among older adults in the United States. Psychoneuroendocrinology. 2023 Mar 1;149:106023.

38. Malatyali A, Sagna De Main A, Cidav T, Komalasari R, Xie R, Thiamwong L. Health Disparities in Cognitive Impairment and Dementia: Role of Social Strain, Depression, and C-Reactive Protein. Gerontol Geriatr Med [Internet]. 2023 Jan 1 [cited 2025 Mar 27];9. Available from: https://journals.sagepub.com/doi/10.1177/23337214231215274

39. De Looze C, McCrory C, O’Halloran A, Polidoro S, Anne Kenny R, Feeney J. Mind versus body: Perceived stress and biological stress are independently related to cognitive decline. Brain Behav Immun. 2024 Jan 1;115:696–704.

40. Christensen R, Miller SP, Gomaa NA. Home-ics: how experiences of the home impact biology and child neurodevelopmental outcomes. Pediatr Res 2024 966 [Internet]. 2024 Sep 28 [cited 2025 Mar 27];96(6):1475–83. Available from: https://www.nature.com/articles/s41390-024-03609-2

41. Gomaa N. Social Epigenomics: Conceptualizations and Considerations for Oral Health. J Dent Res [Internet]. 2022 Oct 1 [cited 2025 Mar 27];101(11):1299–306. Available from: https://journals.sagepub.com/doi/full/10.1177/00220345221110196

42. Karp A, Paillard-Borg S, Wang HX, Silverstein M, Winblad B, Fratiglioni L. Mental, physical and social components in leisure activities equally contribute to decrease dementia risk. Dement Geriatr Cogn Disord [Internet]. 2006 Jan [cited 2025 Mar 20];21(2):65–73. Available from: https://pubmed.ncbi.nlm.nih.gov/16319455/

